# Incidence and Clinical Parameters of HIV-1 Paradoxical and Unmasking Immune Reconstitution Syndrome in Antiretroviral Naïve Pregnant Women attending selected facilities in Kenya

**DOI:** 10.1101/2023.08.22.23294328

**Authors:** John Muthuka K., Everlyn Nyamai M., Oluoch Kelly J., Maibvise Charles, Rosemary Nabaweesi

## Abstract

**Background:** Following ART initiation, a spectrum of HIV-associated immune reconstitution inflammatory syndrome (IRIS), as opportunistic infections occurs, presenting as either paradoxical or unmasking IRIS. There are benefits of ART for maternal health and prevention of perinatal transmission, however, some risks may be possible with IRIS. The study sought to estimate the incidence, compare survival times to, and investigate the baseline clinical predictors of HIV-1 paradoxical and unmasking IRIS as well as identify the homogeneity defined by combinations of specific covariates in Art Naïve Pregnant Women.

**Methods:** A prospective, active records observational study [P609/08/-2018-E] was conducted among ART-naive pregnant women attending antenatal care unit (ANCu) from Kenyatta National and *Mbagathi* Hospitals in Kenya. Participants enrolled were between the ages of 20 - 49 years with a clear documentation of HIV status and, an independent review committee adjudicated IRIS diagnosis. Associations between baseline pre-ART characteristics, biomarkers, and IRIS diagnosis were assessed with Cox models. Decision-tree analysis was finally performed to identify homogeneity defined by combinations of specific parameters.

**Results:** Of the initial total 532 women, (24.8%) (n=133) participants experienced IRIS by the 12^th^ week post ART initiation, 27.3 % (n= 36) and 72.7% (n = 96) paradoxical and unmasking respectively. Maternal Basal Metabolic Index (MBMI) of (25 -29.9) significantly predicted unmasking IRIS [(β)=0.907, Wald test (β^) = 6.550, (HR = 2.478, 95% C.I 1.237 – 4.965, P = 0.010] same case to gravidity of above 5 [(β)=0.743, P = 0.338] while that of 2-3 predicted paradoxical IRIS [(β)= - 0.542, P = 0.037. WHO-HIV clinical stage 1 and 2 showed a link towards paradoxical IRIS, [(β)=- 0.111 and (β)= - 0.276 (P < 0.05)], clinical stage 4 with unmasking IRIS [(β)= 0.047, HR = 1.048, P = 0.941]. CD4 count > 500 cells/mm^3^ skewed towards unmasking IRIS diagnosis [(β)= 0.192, HR = 1.211, P = 0.416], while RNA-HIV viral loads > 50 copies/ml towards paradoxical IRIS [(β)= - 0.199, HR = 0.820, P = 0.360. On decision tree analysis, 76% (P = 0.729) of ART naïve pregnant women aged 20-29 and 40-49 years of age presented with unmasking IRIS and a gravidity of 4-5 and 1 predicted 88% (P = 0.045) unmasking IRIS as compared to that of 2-3 62 % (P = 0.045).

**Conclusion:** Unmasking IRIS was the common type of IRIS post-ART initiation. Maternal BMI and RNA-HIV viral loads level predicted paradoxical IRIS while the baseline CD4 count and WHO-HIV clinical infection stage 1 and 4 were associated with unmasking IRIS. Gravidities of 1 and 4 – 5 predicted unmasking IRIS.

## Background

Immune reconstitution inflammatory syndrome (IRIS) is a clinical complication of some patients infected with the human immunodeficiency virus (HIV) after starting antiretroviral therapy (ART). IRIS is characterized by the release of proinflammatory cytokines and tissue inflammation, and it is related to a well-identified coinfection^1^

All pregnant women with HIV should initiate antiretroviral therapy (ART) as early in pregnancy as possible, regardless of their HIV RNA level or CD4 T lymphocyte count, to maximize their health and prevent perinatal HIV transmission and secondary sexual transmission^2^. One of the goals of antiretroviral therapy (ART) during pregnancy is to achieve and maintain HIV viral suppression to undetectable levels (HIV RNA <50 copies/mL) to reduce the risk of perinatal transmission^2^. Following ART initiation, there occurs a spectrum of HIV-associated IRIS described in several forms of opportunistic infections^3^. This may present as paradoxical or unmasking immune reconstitution inflammatory syndrome (IRIS); “unmasking” IRIS being the flare-up of an underlying, previously undiagnosed infection soon after ART is started while “paradoxical” IRIS being the worsening of a previously treated infection after ART is started^4^.

Despite the known incidence of IRIS in general population^5–8^ and relative knowledge on indirect effect of ART inform of IRIS^9^, the incidence estimation by type of IRIS is not well documented in composite apart from in the nature of opportunistic infections^10–12^. Earlier antiretroviral therapy (ART) initiation and continuation postpartum in PWWH lowers infection rates and mortality, but pregnancy remains an important risk factor for morbidity and mortality even during suppressive ART^13–16^. The physiologic state of pregnancy, independent of HIV, leads to many immunoregulatory changes that help maintain the fetal allograft but may increase susceptibility to infections, including tuberculosis (TB) ^17–20^. A report has demonstrated this increased risk of TB in PWWH, particularly postpartum^21^. Within 6 months of ART, 97 (19.2%) participants developed IRIS and 31 (6.5%) died. A prospective international study established that, in general, persons with lower hemoglobin at baseline were at higher IRIS risk (hazard ratio [HR], 1.2; P =. 004)^22^, a fact that can be relative to other clinical indicators.

Studies have demonstrated higher incidence IRIS among persons living with HIV with a low CD4^+^ T cell count at the baseline^23,24^. Although several aspects of IRIS pathogenesis remain unclear, two clinical parameters have been related to IRIS development: (1) severe CD4+ lymphopenia and (2) opportunistic diseases that lead to a dysregulated immune response (usually characterized by persistent T cell activation, which favors T cell exhaustion)^25,26^. IRIS is diagnosed using the criteria from the International Network for the Study of HIV-associated IRIS (INSHI)^11^. Several studies have focused on identifying biomarkers for IRIS prognosis or diagnosis, but mainly on tuberculosis (TB)-IRIS ^1,27^.

This study was conducted to identify clinical predictors inherent to a specific type of IRIS, either paradoxical or unmasking among as well as looking in to the hazard rates associated with such predictors towards IRIS diagnosis among ART naïve pregnant women. Furthermore, decision tree analysis was conducted to explore the key predictive factors at baseline that can portray a homogenous effect on either paradoxical or unmasking IRIS diagnosis.

## Methods

### Study Design

A prospective, observational and active records study [P609/08/-2018-E] was conducted between May, 2019 and March, 2020. HIV-positive confirmed, ART-naive pregnant women attending antenatal care unit (ANCu) from the first trimester and not later than one-month post-conception. The cohort was then initiated to ART as a single population at baseline. IRIS as the primary outcome was assessed and confirmed within the first 12 weeks using International Network for the Study of HIV-associated IRIS (INSHI) and experts’ opinions as per IRIS case definitions was done to ensure internal validity was maintained, while secondary outcome was any form of adverse pregnancy outcome (APO) from the end of first trimester to post-delivery period.

Baseline clinical and demographic characteristics were recorded among these of pregnant women visiting at 2 health facilities in Kenya, namely, Kenyatta National and *Mbagathi* Hospitals. Inclusion criteria were age ≥20 years, documentation of HIV status, ART naïve, basal metabolic index, CD4 in cells/µL, and HIV-RNA (log10 copies/mL). Exclusion criteria were known critical conditions and any severe threatening events in pregnancy. Consecutive eligible pregnant women were enrolled including the utilization of their inherent active records. The protocol was approved by the ethics committees of the participating facilities. Participants were followed through their active records prospectively from ART initiation (week 0) to week 12. ART regimens and timing of initiation were chosen according to local treatment guidelines and clinicians’ preferences. The primary objective was to estimate cumulative incidence of IRIS as well as by the type of IRIS (either unmasking or paradoxical) by week 12.

The clinical teams at study sites prospectively identified IRIS events and collected relevant clinical information that was presented to an endpoint review committee. The committee determined whether the events were consistent with IRIS using the following AIDS Clinical Trials Group IRIS definition criteria ^28^. Evidence of ART initiation with resultant increase in CD4 count (≥50 cells/µL or a ≥2-fold rise) and/or virologic suppression (>0.5 log10 decrease in plasma HIV viremia), clinical presentation consistent with an infectious or inflammatory condition, and the absence of an alternative etiology such as the expected course of a previously recognized infection or side effects of medications.

### Statistical Methods

Descriptive univariate analysis considering IRIS cumulative incidence grouped by IRIS type was performed not ignoring censoring and to investigate the relationship between clinical characteristics with the type of IRIS among participants within the first phase of the study at 12 weeks after starting ART, a bivariate analysis was performed using Pearson-test for clinical parameters relative to IRIS type. For the baseline clinical indicators and time to a specific type of IRIS, multivariate Cox regression analysis with IRIS as the outcome was fitted, with censoring effect included prior to 12 weeks post ART initiation, using the *Survival* package in SPSS ^29^. To explore further key relationship between baseline characteristics and IRIS type diagnosis, a decision tree analysis was performed.

## Results

A total of 532 HIV-infected, ART-naive pregnant women participants were screened with 143 diagnosed with IRIS recruited, of which 132 remained after exclusion of eleven (n=11) following lack of consensus on IRIS presentation at phase one of the study in the first trimester as well as other critical conditions. The 132 participants were enrolled at the 2 sites (90 Kenyatta National Hospital, and 41 in *Mbagathi* between August 2019 to May 2020. Majority of women (53%, n=70) were between the ages of 30-39, 51% (n = 69, had a normal body mass index (18.5-24.9), and 80 (60%) were married. Most, 50% (n= 67) had a parity of 2-3. Over 50% (n = 73), were at the first stage of HIV infection as determined in WHO guidelines^30^ and 5.3% (n = 7) had maternal anemia with 42% (n = 56) presenting with at least a symptom of an opportunistic infection.. Tenofovir alafenamide/emtricitabine (TAF/FTC) combination was started in 72% (n = 96) of the pregnant women initiated on ART while few, 5.3% (n=11) were put on tenofovir disoproxil fumarate/emtricitabine (TDF/FTC; *table 1*.

**Table 1:** Baseline characteristics of ART naïve pregnant women at the first trimester.

| Variable |  | Frequency | Percent | (%) |
| --- | --- | --- | --- | --- |
| **WHO-HIV Stage** | Primary | 36 | 17.2 | 27.1 |
|  | Stage 1 | 73 | 34.9 | 54.9 |
|  | Stage 2 | 21 | 10.0 | 15.8 |
|  | Stage 3 | 3 | 1.4 | 2.3 |
| **Opportunistic Infections** | Present | 56 | 26.8 | 42.4 |
|  | Absent | 76 | 36.4 | 57.6 |
| **Maternal age (Years)** | 20-29 | 40 | 19.1 | 30.1 |
|  | 30-39 | 70 | 33.5 | 52.6 |
|  | 40-49 | 23 | 11.0 | 17.3 |
| **ART Combination** | (ABC/3TC) * | 26 | 12.4 | 19.5 |
|  | (TAF/FTC) | 96 | 12.4 | 72.2 |
|  | (TDF/FTC) * | 11 | 5.3 | 8.3 |
| **Maternal age** | 20-29 | 40 | 19.1 | 30.1 |
|  | 30-39 | 70 | 33.5 | 52.6 |
|  | 40-49 | 23 | 11.0 | 17.3 |
| **MBMI** | (< 18.5) | 21 | 10.0 | 15.8 |
|  | (18.5-24.9) | 69 | 33.0 | 51.9 |
|  | (25 -29.9) | 35 | 16.7 | 26.3 |
|  | (> 30) | 8 | 3.8 | 6.0 |
| **Maternal anemia** | Present | 7 | 3.3 | 5.3 |
|  | Absent | 126 | 60.3 | 94.7 |
| **Rhesus Factor** | Positive | 116 | 55.5 | 87.2 |
|  | Negative | 17 | 8.1 | 12.8 |
| **Parity** | 1 | 50 | 23.9 | 37.6 |
|  | 2-3 | 67 | 32.1 | 50.4 |
|  | 4-5 | 14 | 6.7 | 10.5 |
|  | >% | 2 | 1.0 | 1.5 |
| **Marital Status** | Single | 3 | 1.4 | 2.3 |
|  | Separated | 41 | 30.8 | 30.8 |
|  | Married | 80 | 60.2 | 60.2 |
|  | Windowed | 8 | 3.8 | 6 |

\*Abacavir/lamivudine (ABC/3TC) or either tenofovir alafenamide/emtricitabine (TAF/FTC) or tenofovir disoproxil fumarate/emtricitabine (TDF/FTC)

One hundred and thirty-two, (24.8%) participants experienced IRIS by the 12^th^ week post ART initiation. The proportions of IRIS by type were 27.3 % (n= 36) and 72.7% (n = 96) paradoxical and unmasking respectively; *table 2*.

**Table 2:**
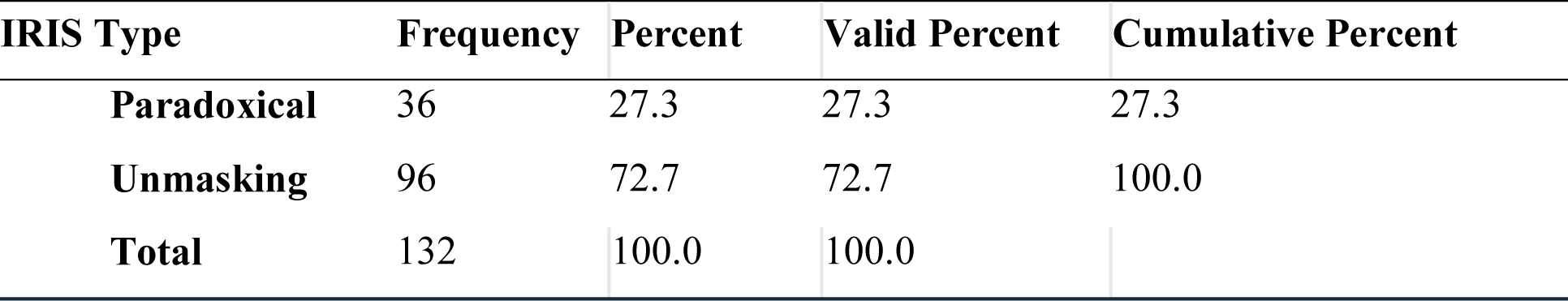
Cumulative incidence of paradoxical and unmasking IRIS.

| IRIS Type | Frequency | Percent | Valid Percent | Cumulative Percent |
| --- | --- | --- | --- | --- |
| **Paradoxical** | 36 | 27.3 | 27.3 | 27.3 |
| **Unmasking** | 96 | 72.7 | 72.7 | 100.0 |
| **Total** | 132 | 100.0 | 100.0 |  |

### Survival and hazard function at the mean of baseline clinical covariates

The total accumulated risk of experiencing IRIS symptoms as demonstrated by the cumulative hazard function evaluating all the clinical parameters increased with time more so from week 8 onwards to 12^th^ week; *figure 1*.

**Figure 1:**
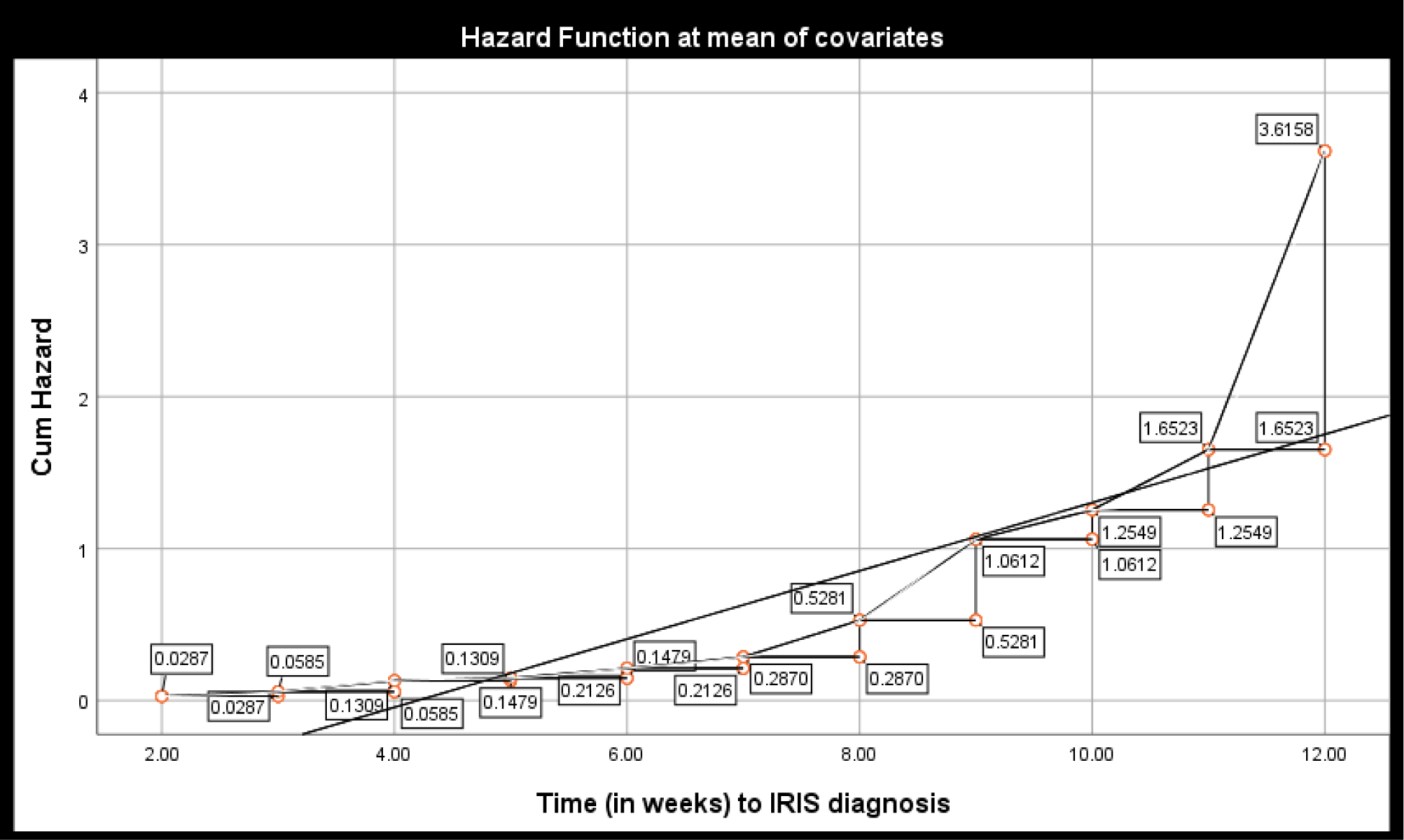
Cumulative hazard for IRIS in general as shown by hazard functions for the clinical parameters.

### Clinical parameters at baseline and association with paradoxical or unmasking IRIS

Clinical parameters at baseline following Cox-regression analysis demonstrated that, maternal Basal Metabolic Index (MBMI) of (25 -29.9) was statistically significant for unmasking IRIS as opposed to paradoxical IRIS with a positive regression coefficient [(β)=0.907, Wald test (β^) = 6.550, (HR = 2.478, 95% C.I 1.237 – 4.965, P = 0.010]. Seemingly, although insignificant, MBMI > 30 had positive regression coefficient towards unmasking IRIS [(β)=0.935, P = 0.122]. In general, HR for IRIS increased with an increase in BMI levels; *figure 2*.

**Figure 2:**
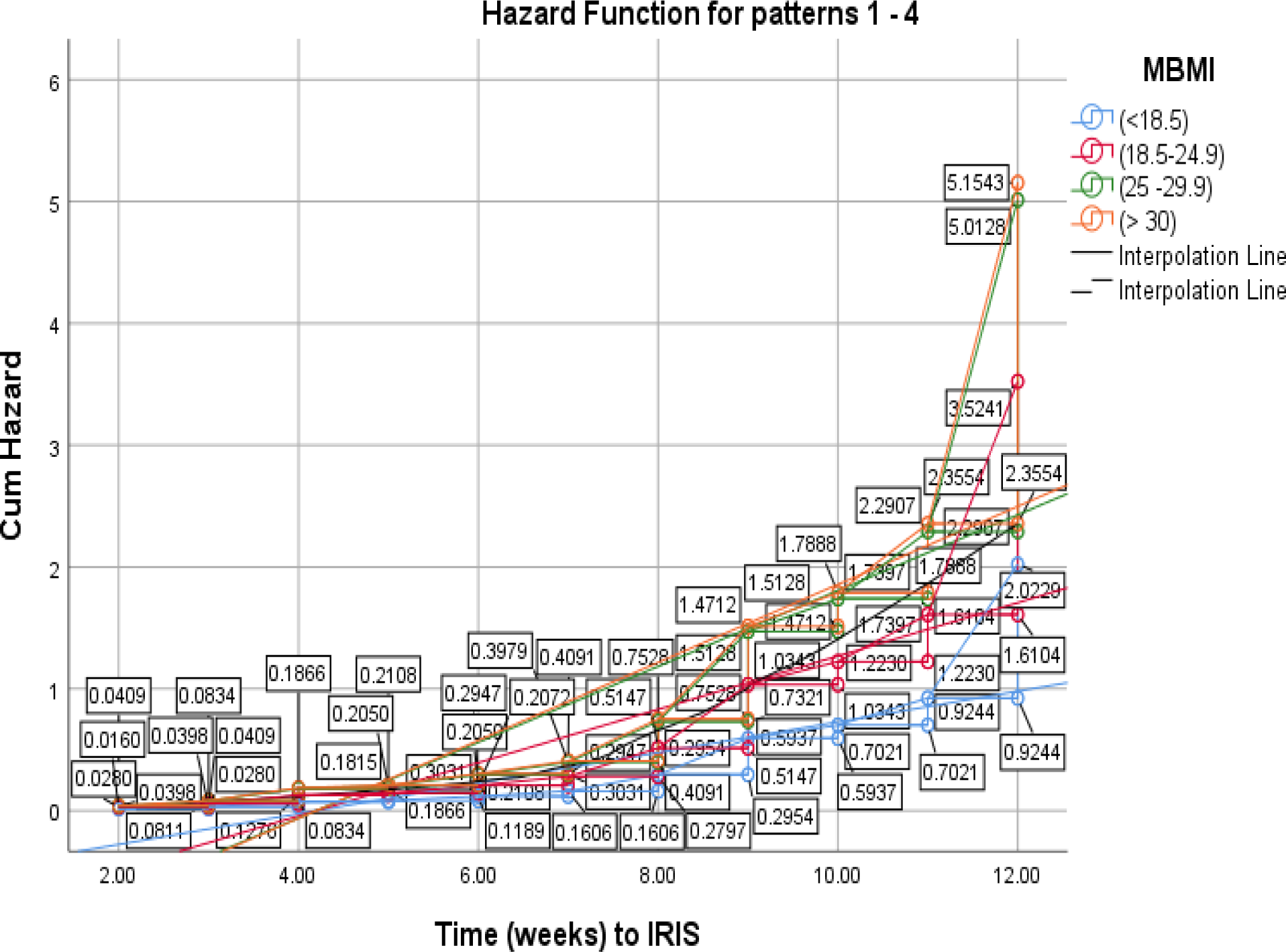
Hazard function for IRIS diagnosis by maternal basal metabolic index (MBMI)

Similarly, a parity of above 5 [(β)=0.743, P = 0.338] as opposed to those with a parity of below 5; parity of 2-3 [(β)= -0.542, P = 0.037] clearly predicted paradoxical IRIS as well as a parity of 3-4, however, statistically insignificant [(β)= - 0.163, P = 0.850]. WHO-HIV Staging (clinical stage 1) and (clinical stage 2) showed a negative regression coefficient (β)= - 0.111 and (β)= - 0.276 (P < 0.05) respectively, a link towards paradoxical IRIS although statistically insignificant. WHO-HIV Staging (clinical stage 4) was positive with unmasking IRIS, statistically insignificant [(β)= 0.047, HR = 1.048, P = 0.941]. The HR for unmasking IRIS was higher at primary and clinical stage 4 both with a hazard function of above 4.0 at the end of 12 weeks as opposed to other stages; *figure 3*.

**Figure 3:**
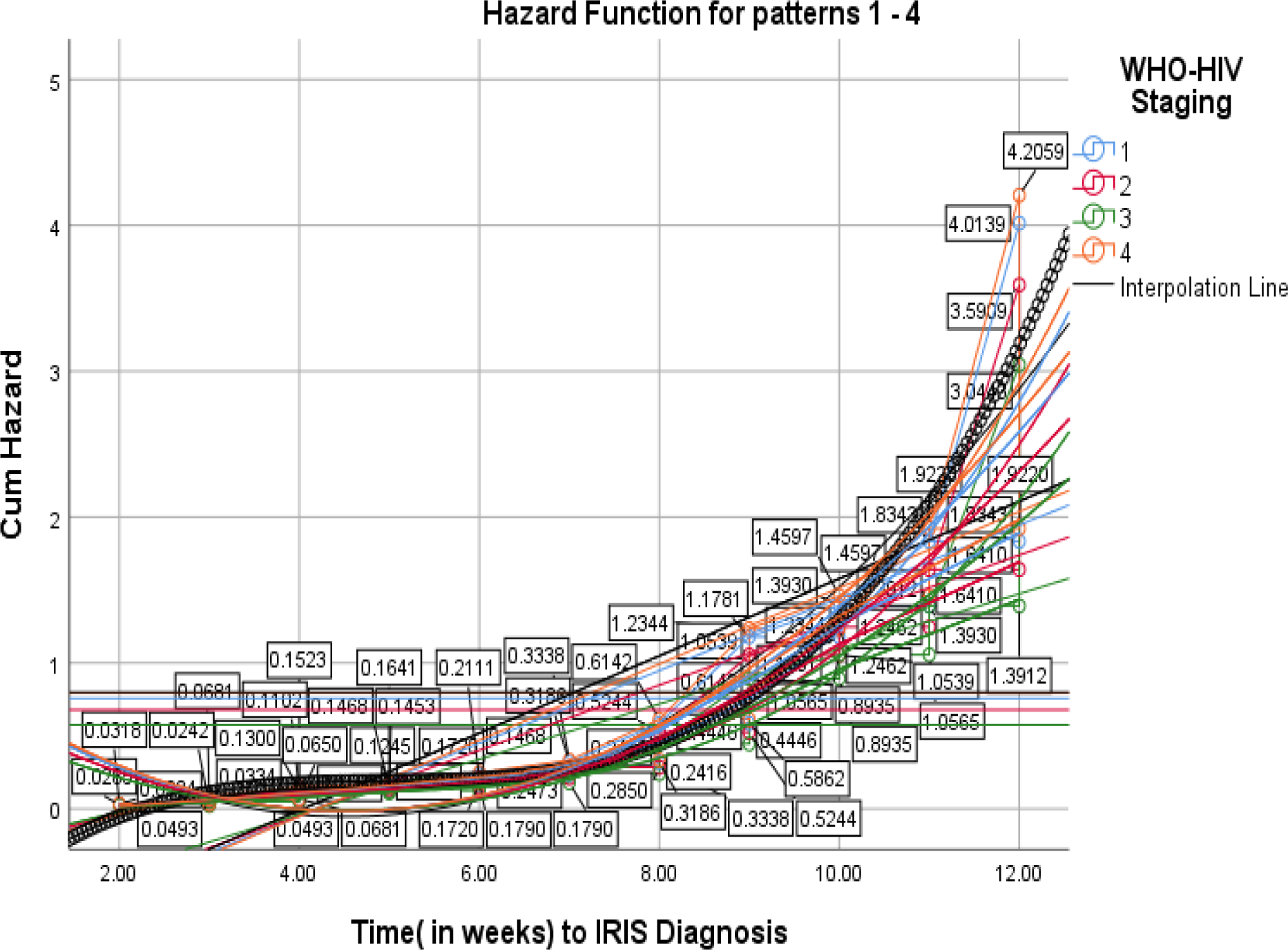
Hazard function for IRIS diagnosis by WHO-HIV Staging.

CD4 count of over 500 cells/mm^3^ seemingly was associated with unmasking IRIS diagnosis [(β)= 0.192, HR = 1.211, P = 0.416]; *figure 7,* while RNA-HIV viral loads above 50 copies/ml with paradoxical IRIS [(β)= - 0.199, HR = 0.820, P = 0.360]. The cumulative hazard for IRIS in general was higher in pregnant women having CD4 count > 500cells/mm^3^ and HIV viral loads> 50 Copies /mL at baseline and this risk, increased with time towards the 12^th^ week post ART initiation; *figures 4 and 5* respectively.

**Figure 4:**
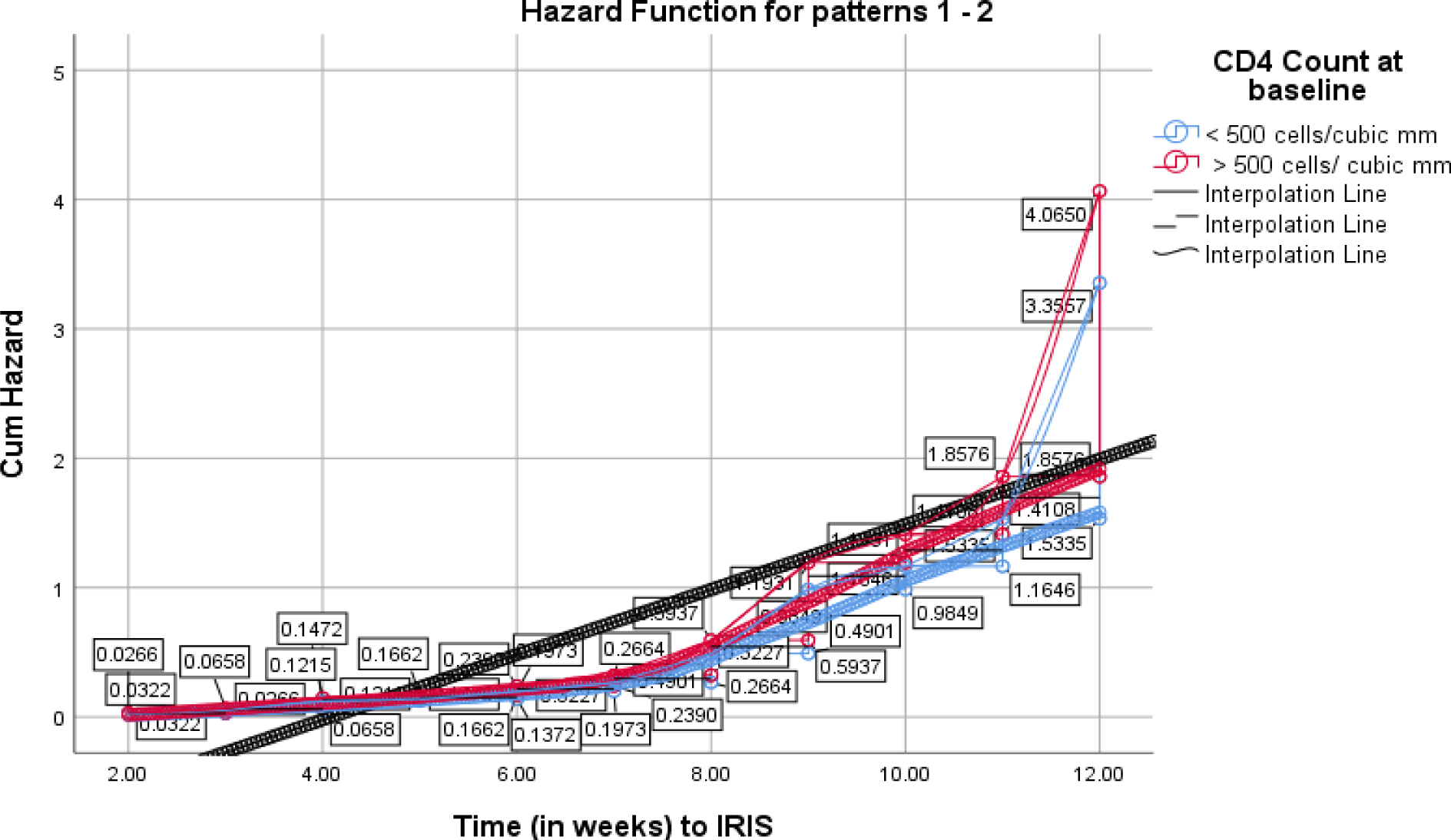
Hazard function for IRIS diagnosis by CD4 count at baseline.

**Figure 5:**
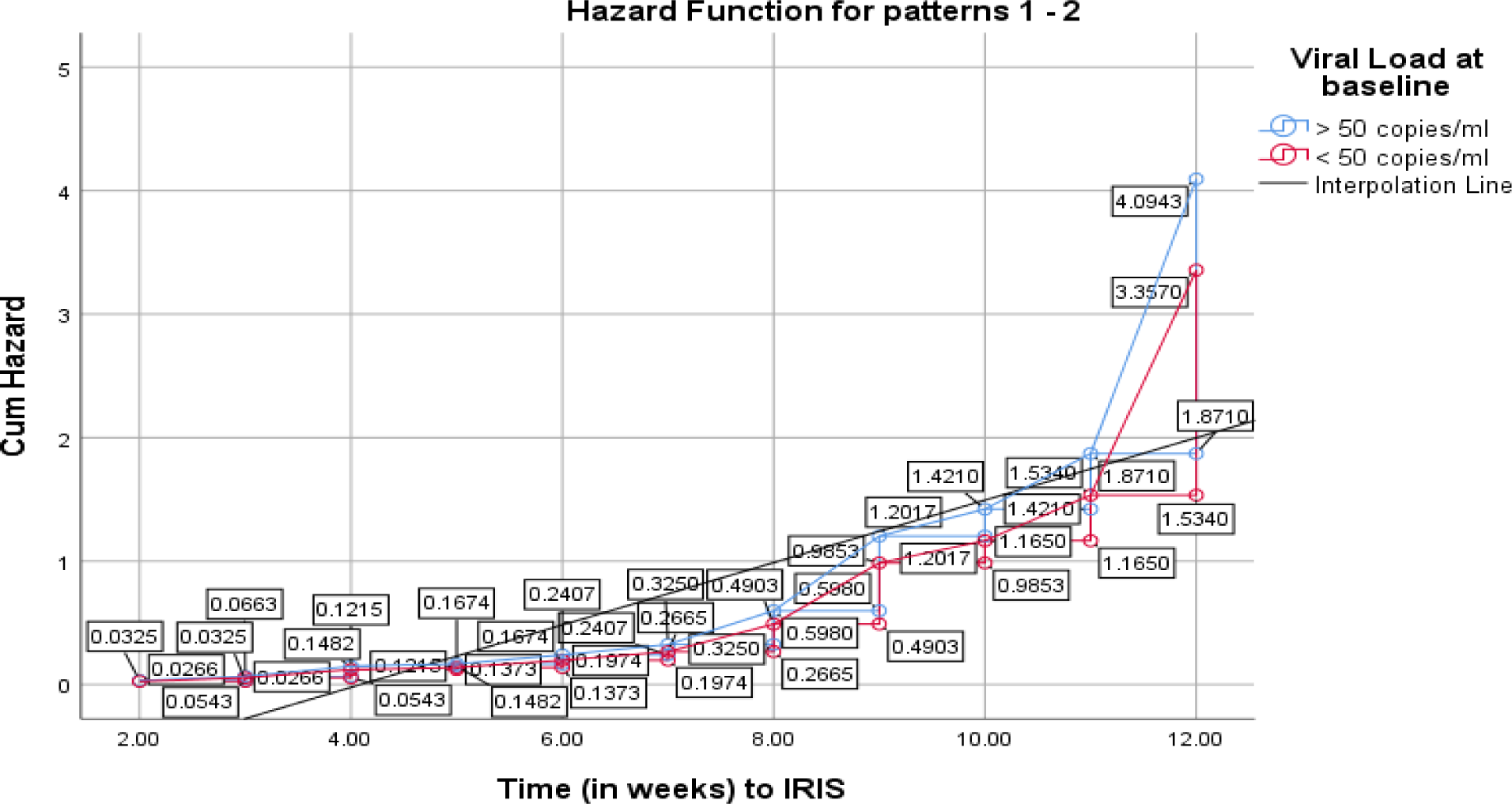
Hazard function for IRIS diagnosis by RNA-viral loads at baseline.

All the above specific parameters are herein is presented; *table 3*

**Table 3:**
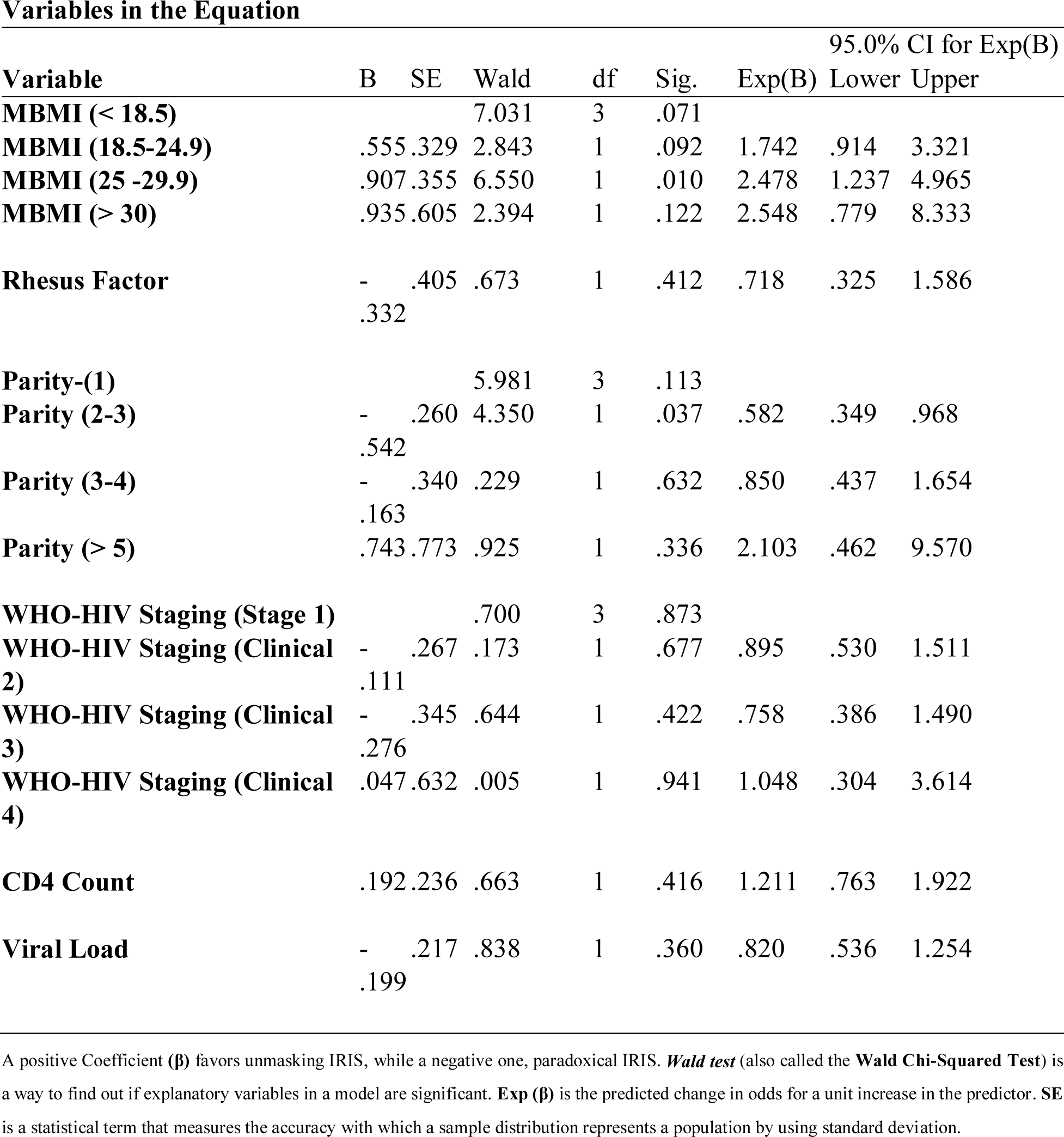
Cox-model regression analysis for clinical parameters at baseline.

### Decision tree analysis on prediction of IRIS incidence

Collectively for exploratory and confirmatory classification analysis to assess their predictions of IRIS type by 12^th^ week post ART initiation, decision tree analysis was performed by including all the clinical parameters and the demographic characteristics of the participants including; location, education, occupation, religion, marital status, income source and maternal age. Maternal age was the only notable predictor of IRIS type. Seventy six percent (76%) (P = 0.729) of pregnant women aged 20-29 and 40-49 years of age were diagnosed with unmasking IRIS. This prediction applied to 63 women and the model was accurate 48 times. Further, women with a gravidity of 4-5 and 1 predicted eighty-eight (88%) (P = 0.045) unmasking IRIS as compared to those with a parity of 2-3 (62 %) (P = 0.045); *Table 4*.

**Table 4:**
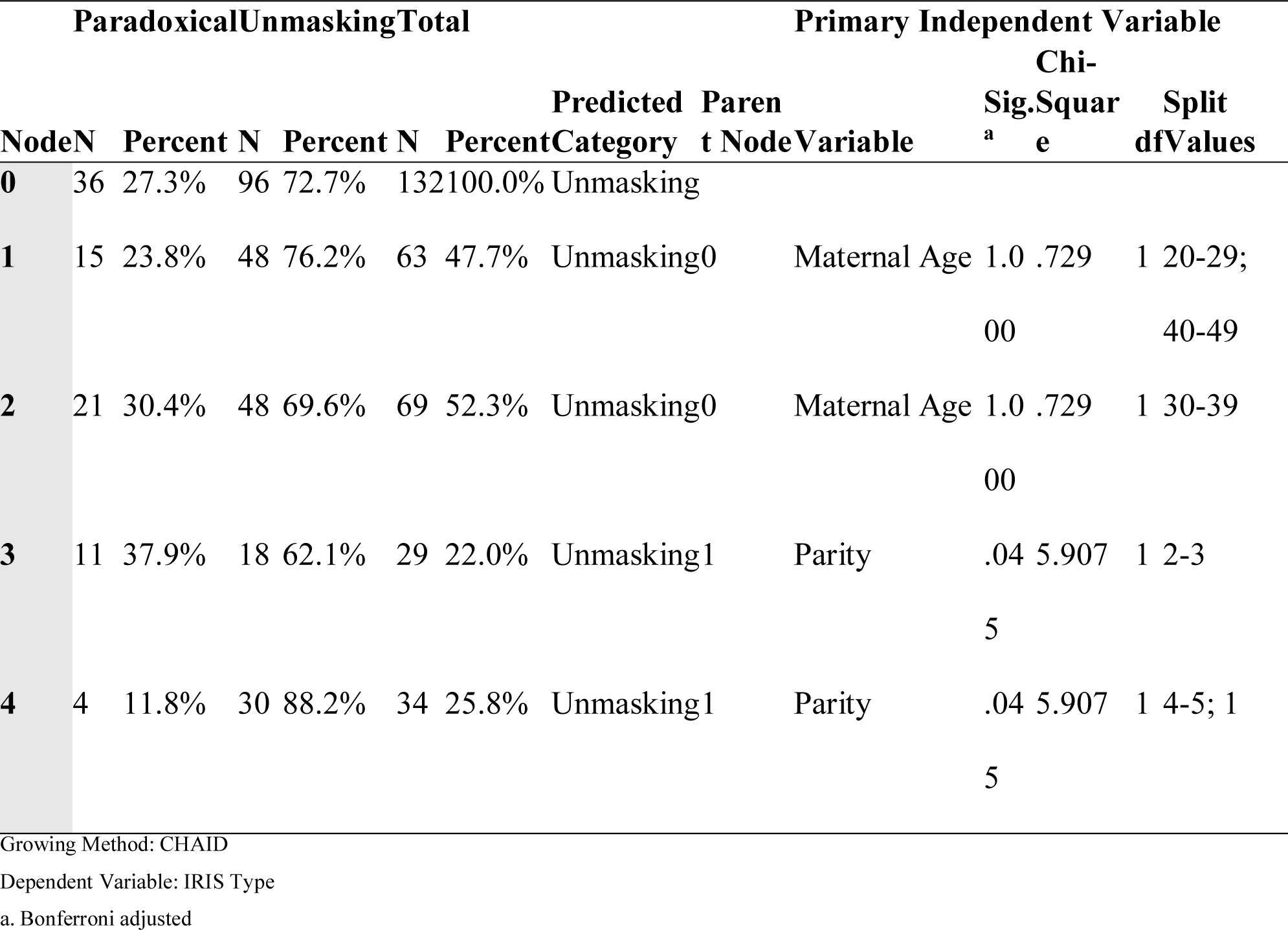
Tree table on exploratory decision tree analysis of IRIS development by pooled baseline characteristics.

The gain chart; figure 6 demonstrated that unmasking IRIS was predicted close to the perfect prediction model with the ratio of the node response percentage for the target category compared to the overall target category response percentage for the entire sample; *figure 7*.

**Figure 6:**
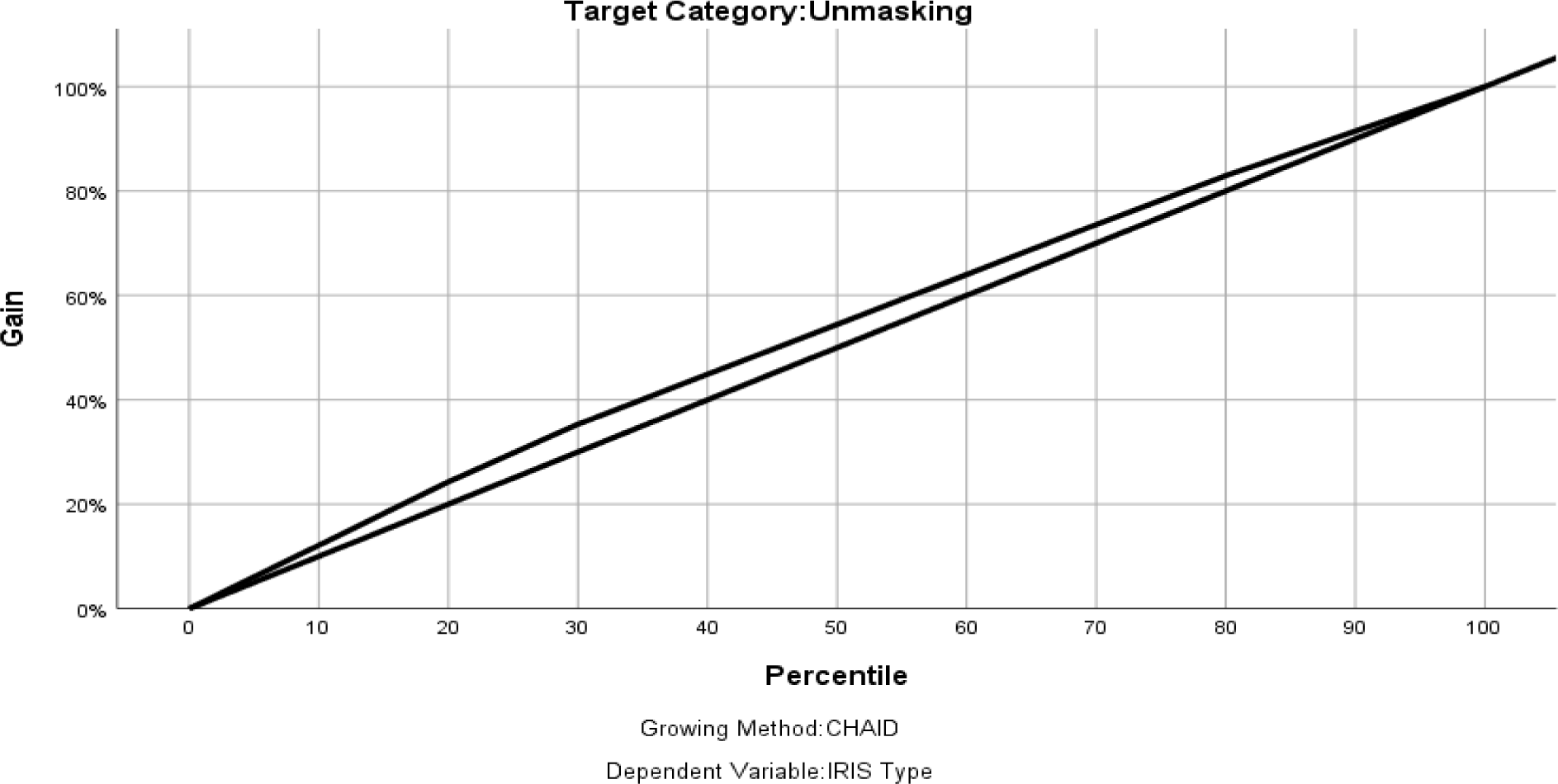
The gain chart curve on prediction model of IRIS.

**Figure 7:**
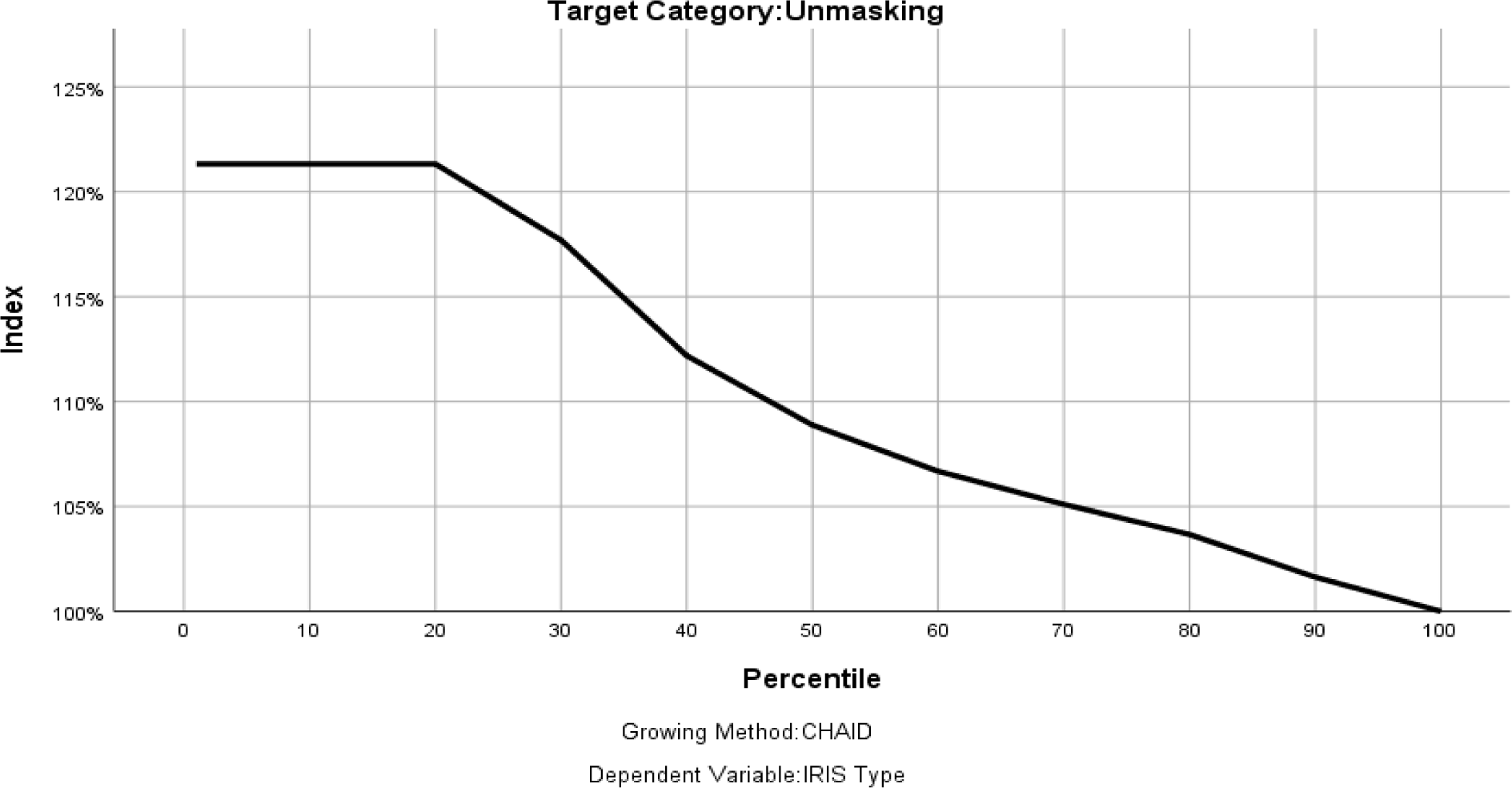
The index chart on the ratio of the node response percentage.

## Discussion

In this prospective, observational, active records-based study conducted in 2 facilities, representing diverse clinical setting and environments in which IRIS may occur, we evaluated IRIS incidence by type and IRIS predictors including survival and hazard risk analysis using Cox-model.

Unmasking IRIS was more common in this population of ART naïve pregnant women as compared to paradoxical IRIS similar to a study that reported 69.4% “unmasking” and 27.8% “paradoxical”^31^. This was further close within the percentage range of 59% to 76% obtained from other studies ^27,32–36^. Further clearly as demonstrated in this current, unmasking verses paradoxical IRIS were found to be (69.4%) “unmasking” and 27.8% “paradoxical”^12^. Another study, reports a 22% paradoxical IRIS ^37^, close to the current proportion as obtained from this study.

Maternal Basal Metabolic Index (MBMI) of (25 -29.9) clearly predicted unmasking IRIS as depicted is a study that established BMI ≥20 kg/m^2^ as probable predictor of IRIS ^27,38^, contrary to another study that established BMI <18.5 (HR 2.15, 95% CI 1.07–4.3, p= 0.03) as IRIS diagnosis predictor. Gravidity of above 5 clearly predicted unmasking IRIS and although there are no clear studies elucidating this fact, a study concluded that, gestational age, and gravidity of above 4 influenced T lymphocyte subset levels ^39^, a key phenomenon in IRIS development. WHO-HIV Staging (clinical stage 4) was positive with unmasking IRIS, and as it was the common form of IRIS, this findings are elucidated in a studies that report HIV late presenters ^40,41^. Again, another study shows risk factors for IRIS include an advanced state of immunosuppression disseminated OIs at ART initiation^32,42^, a clear fact associated with advanced HIV infection The hazard risk for unmasking IRIS was higher at primary and clinical stage 4. This is matched in a study that described five patients with advanced HIV-1 infection in whom initiation of HAART resulted in unmasking of an underlying occult opportunistic infection^43^, and as regards unmasking IRIS diagnosis, this can be attributed to the fact that, people with acute HIV infection may experience fever, lymphadenopathy, pharyngitis, skin rash, myalgia, arthralgia, and other symptoms^44–46^.

In contrast to other studies that showed higher risk IRIS in people starting treatment with a very low CD4 cell count (usually below 100) ^6,42,47–49^, the current study established that, CD4 count of over 500 cells/mm^3^ seemingly was associated with unmasking IRIS diagnosis in this ART naïve cohort of pregnant women. This fact may be due to pregnancy related phenomenon and further, IRIS a pathologic inflammatory response to a preexisting antigen that develops soon after initiation of ART therapy with or without a corresponding increase in CD4 cell count ^50^. Again, the already high CD4 count prior to ART initiation may imitate the known increase of the same that is associated with developing IRIS^51,52^. Further to this, IRIS can occur IRIS can occur at any CD4 count 5,8,53–55

On the other hand, RNA-HIV viral loads > 50 copies/ml showed a positive direction with paradoxical IRIS, similar to existing evidence ^55–58^. However, on contrary, some reports have shown that, it is the dramatic reduction of HIV-RNA viral load levels after starting ART that significantly demonstrate IRIS development^55,59,60^. This concept may be justified by the evidence that, pre-existing latent opportunistic infection with a high antigenic burden at the time of starting ART increases the risk and severity of IRIS^8,37,55,61^. In real sense, paradoxical IRIS is linked with most opportunistic infection as compared to unmasking IRIS^27,62,63^. The cumulative hazard for IRIS was higher in pregnant women with both higher baseline CD4 count and viral loads increasing with time towards the 12^th^ week post ART initiation. This, concurs with findings that showed increased baseline CD4+ T-cell count among women of reproductive age on ART ^64^, while for higher viral loads by other studies^65^. The increased risk for IRIS with time as per this study further is generally implicated in previous studies ^47,66^. Decision tree analysis showed that, majority of much younger and older women presented with unmasking IRIS and seemingly, a gravidity of 1 and 4-5 were associated with unmasking IRIS. This finding is supported by a study conducted in southern Mozambique^67^.

This study had several limitations. Identification of IRIS by type, either paradoxical or unmasking may have been compromised due to the thin gap between the two, however, the clear approach was based and guided by the specific biomarkers such as CD4 count and RNA-viral loads at baseline. Again, the duration of time a woman had stayed after contracting HIV may not have been known prior to conceiving or after, however, based on the clinical parameters guiding the HIV infection staging by WHO, it was possible to estimate the same. Last, a pregnant woman, due to a slow developing disease courtesy of having been infected with HIV, may have used some medications without clinical directives that would have compromised the process of IRIS identification. This however was tackled by using a combined guideline to diagnose IRIS and as such, this limitation somehow mitigated. Despite these limitations, we believe the study results provide valuable information about the incidence of IRIS by type, either unmasking or paradoxical., estimates the survival time to the event of either IRIS in general and IRIS by type while accounting for key baseline clinical parameters predicting IRIS in ART naïve pregnant women.

In conclusion, as per this study, unmasking IRIS was the common type of IRIS. BMI of 25-29 and baseline viral loads of 50 copies /ml were associated with paradoxical IRIS while the baseline CD4 count of 500 cells/ mm^3^ with unmasking IRIS, same case with WHO-HIV clinical infection stage 1 and 4. Gravidities of 1 and 4 – 5 were predicted unmasking IRIS. Future prospective studies should further validate the potential predictive value, feasibility, and effectiveness of BMI, gravidity, age, gestational stage, HIV baseline biomarkers and HIV infection clinal sage in predicting unmasking or paradoxical IRIS, and further, harness the evidence on survival time to an occurrence of IRIS among ART by type naïve pregnant women.

## Data Availability

All data produced in the present study are available upon reasonable request to the authors

